# The Alpha-1 Antitrypsin Z-allele associates with sarcopenia in the UK Biobank

**DOI:** 10.64898/2026.09.12.26362905

**Authors:** Amy Attaway, Karina Serban, Shovan Dutta, Umur Hatipoğlu, Joe Zein, Peter Bazeley, Maria Kyriakidou, Jennifer E. Dawson, Lisa Ruvuna, R. Chad Wade, Vickram Tejwani, Jessica Bon, J. Michael Wells, Victor Ortega, James K. Stoller, Russell Bowler

**Affiliations:** Departments of Pulmonary Medicine, Cleveland Clinic, Cleveland, Ohio; Genomic Sciences and Systems Biology, Cleveland Clinic, Cleveland, Ohio; Departments of Medicine/ Division of Pulmonary, Critical Care, Sleep Medicine and Molecular Genetics and Microbiology, University of Florida. Gainesville, FL; Critical Care, Cleveland Clinic, Cleveland, Ohio; Division of Pulmonary Medicine, Department of Medicine. Mayo Clinic Arizona. Scottsdale, AZ; Quantitative Health Sciences, Cleveland Clinic, Cleveland, Ohio; Section of Pulmonary, Critical Care, Allergy, and Immunologic Diseases, Wake Forest University School of Medicine. Winston-Salem, NC; Division of Pulmonary, Allergy, and Critical Care Medicine, University of Alabama at Birmingham (UAB). Birmingham, AL; Education Institute, Cleveland Clinic, Cleveland, Ohio

**Author notes:** Denotes co-senior author. Data access and analyses were performed following approval by the Institutional Review Board (IRB #20-446).

**Keywords:** Alpha-1 antitrypsin deficiency, sarcopenia, COPD, emphysema

## Abstract

**Background:** Sarcopenia is common in chronic obstructive pulmonary disease (COPD) and contributes to adverse outcomes. Alpha-1 antitrypsin deficiency (AATD) is the most common genetic cause of COPD, but its impact on skeletal muscle remains to be determined.

**Methods:** We performed a retrospective analysis of 487,207 participants in the UK Biobank with available *SERPINA1* genotyping. Associations between AAT genotypes and measures of skeletal muscle mass were determined using multivariable linear and logistic regression models. Models adjusted for age, sex, smoking status, genetic principal components, lung function (FEV₁% predicted), and liver fibrosis risk (FIB-4). We determined that the Z-allele was associated with sarcopenia and mortality using regression models and mediation analysis.

**Results:** Compared with PI*MM individuals, carriers of the Z allele (PI*MZ, PI*SZ, PI*ZZ) demonstrated significantly lower fat free mass index (FFMI) and appendicular skeletal muscle index (ASMI), as well as higher odds of sarcopenia by binary definition. Associations persisted after adjustment for lung and liver function. A dose-dependent relationship was observed, with the greatest reductions in muscle mass and highest sarcopenia associated with PI*ZZ individuals (adjusted OR for FFMI-defined sarcopenia 2.02, 95% CI 1.16-3.32). Each additional Z allele was associated with increased mortality risk (adjusted OR 1.09, 95% CI 1.02-1.16), with sarcopenia mediating a modest proportion (∼2.6%) of this association.

**Conclusions.:** The *SERPINA1* Z allele is associated with reduced muscle mass and increased risk for sarcopenia independent of lung and liver disease. Our findings expand the phenotypic spectrum of AATD-related diseases to include skeletal muscle as a modifiable target for intervention.

## Introduction

Sarcopenia, i.e., the loss of skeletal muscle mass or function, occurs in up to 50% of patients with chronic obstructive pulmonary disease (COPD) and is a major contributor to morbidity, mortality, and healthcare utilization(1). Alpha-1 antitrypsin deficiency (AATD), the most common genetic cause of COPD, is due to variation in the *SERPINA1* locus. Severe deficiency causes emphysema due to loss of AAT protein function and causes liver disease due to misfolded protein accumulation(2). The protease inhibitor (PI) Z genotype (Glu342Lys) accounts for 95% of AATD-related lung disease and causes a structural change to the AAT protein that predisposes to polymerization, leading to intracellular PI*Z-polymer accumulation and proteotoxic stress that cause liver disease(3). Beyond hepatocyte accumulation, PI*Z-polymers are secreted into the circulation, contribute to systemic inflammation(4), and have been shown to accumulate in other organs including adipose(5). In addition to the PI*ZZ genotype, other AAT genotypes pose disease risk. The compound heterozygote genotype PI*SZ, where the PI*Z-allele is coinherited with another coding variant, PI*S (Val264Glu), is associated with an increased risk of emphysema amongst smokers(6). Recent literature has also shown that PI*MZ heterozygotes have a low absolute risk of lung disease, but that this risk increases substantially among those with significant smoking exposure(7).

Prior work analyzing the UK Biobank (UKBB) and the Nationwide Inpatient Sample (NIS) shows that sarcopenia is common in COPD and is associated with increased mortality(1), hospitalizations, and higher healthcare utilization(8). While COPD and liver disease are common complications of AATD, recently published work in the NIS(9) suggest an independent risk of sarcopenia in patients with AATD. However, because the NIS relies on insurance billing codes, AATD diagnosis is dependent on administrative coding which may be inaccurate (12) and cannot distinguish between specific AATD genotypes. Because genotype confirmation is the definitive method for diagnosing and risk-stratifying AATD, it remains unknown whether the increased sarcopenia risk is specific to individuals carrying the Z-allele. The NIS also does not provide granular data on muscle mass, and therefore the degree of muscle loss associated with AATD is also unknown.

We hypothesize that the prevalence of sarcopenia is greater in those with AATD, particularly those carrying a PI*Z allele, compared to those without AATD. Further, we hypothesize that the association between sarcopenia and the PI*Z-allele is independent of liver or lung disease. To test these hypotheses, we analyzed the UKBB to assess PI*Z genotype-specific effects on skeletal muscle mass and body composition, and to determine whether sarcopenia amongst individuals with PI*Z variants is associated with increased risk of mortality.

## Methods

### Study Population and Data Source

We retrospectively analyzed baseline UK Biobank (UKBB) data from adults enrolled 2006–2010, including 487,207 participants of European descent (**Supp Fig 1**) with non-missing SERPINA1 Z (rs28929474) and S (rs17580) genotypes. Both variants are directly genotyped, obviating the need for imputation. Minor allele frequencies were 0.020 (Z) and 0.048 (S). In accordance with UKBB data-sharing policy, clinical characteristics fewer than 10 participants were not reported to protect participant privacy and reduce the risk of re-identification. Comorbidity definitions and phenotyping are detailed in the Supplementary Methods.

**Figure 1.**
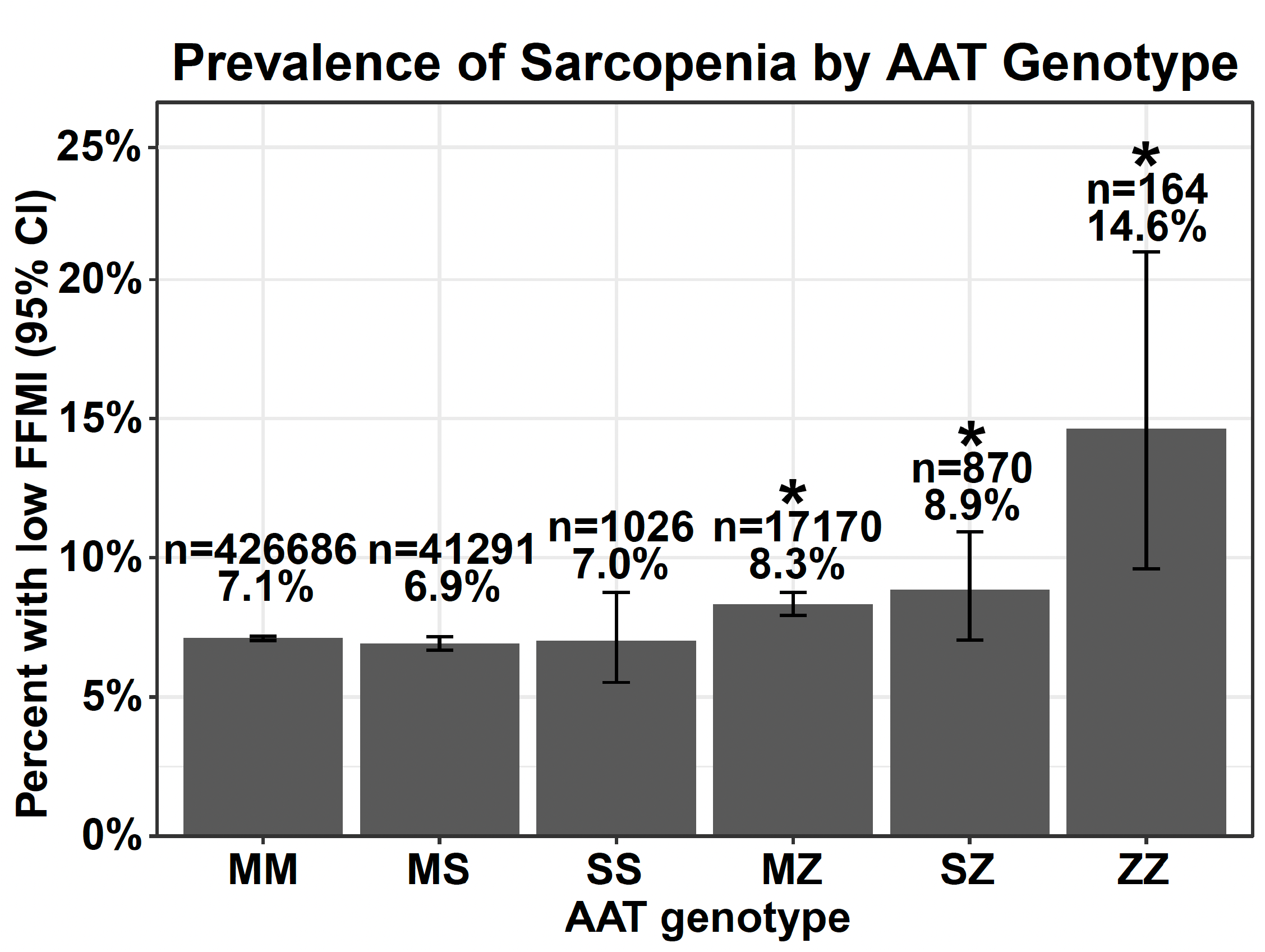
Prevalence of FFMI-defined sarcopenia across AAT genotypes. FFMI-defined sarcopenia prevalence demonstrated a genotype-dependent pattern, with progressively higher proportions observed among Z allele carriers and the highest prevalence in ZZ individuals (14.6%). Error bars represent 95% confidence intervals. Sample sizes are indicated above each bar. * indicates statistically significant prevalence by Z test of proportions compared to PI*MM.

### Descriptive Statistics

Normally distributed continuous measures were summarized as means (SD) and compared by AAT genotype; categorical variables were compared using Pearson’s chi-square tests.

### Muscle Mass Assessment

Fat-free mass index (FFMI = fat-free mass/height²) and appendicular skeletal muscle mass index (ASMI = appendicular lean mass/height²) are validated measures of skeletal muscle mass in chronic disease populations including COPD. Body composition was measured by bioelectrical impedance (Tanita BC-418 MA; Tanita Europe). Sarcopenia was defined by low FFMI (<17.4 kg/m² for males and <15.0 kg/m² for females(10)) or low ASMI (<8.90 kg/m² in males and <6.37 kg/m² in females(11)), consistent with established thresholds. Handgrip strength (HGS) was assessed using a Jamar J00105 hydraulic dynamometer, with low HGS defined as <30 kg for males and <20 kg for females(12).

### Phenome-Wide Association Study and Data Visualization

A PheWAS plot visualized associations between Z allele dosage and all UKBB phenotypes. Pre-specified phenotypes were classified using keyword-based pattern matching into three groups: “Known Alpha-1 Associated Phenotypes” (e.g., emphysema, bronchitis, bronchiectasis, FEV1, FVC, liver parameters), “Muscle/body composition” (e.g., fat free mass, grip strength, physical activity, bone parameters), or “Other”. Phenotypes were ranked by descending −log₁₀(p), with rank as the x-axis position and a reference line at Bonferroni significance (−log₁₀ > 1.3).

### Regression Models

Continuous outcomes (FFMI, ASMI) were modeled with multivariable linear regression and binary sarcopenia with multivariable logistic regression, reported as β coefficients or odds ratios with 95% CIs. Primary outcomes were FFMI (continuous) and FFMI-defined sarcopenia; secondary outcomes were the ASMI equivalents.

### Covariable Selection

Covariables associated with AAT were chosen *a priori* based on our review of literature and included sex, age at assessment, and smoking status (current vs. former vs. never)(7, 13). Pack-year history was not included in primary models due to substantial missingness (>70% missing). BMI was not included, as it shares components with FFMI and ASMI (BMI = [fat-free mass + fat mass]/height²), raising concern for over-adjustment bias(14, 15). Genetic principal components 1–5 were included given a small but significant association with muscle outcomes (R²=0.003). Ethnicity was not included because certain genotypes (e.g., PI*ZZ) were represented entirely by White participants, consistent with the known ancestral distribution of AAT deficiency alleles(16). To test independence from pulmonary and hepatic dysfunction, fully adjusted models added FEV1% predicted (GLI reference)(17) and the Fibrosis-4 index (FIB4), a validated non-invasive score derived from age, AST, ALT, and platelet count(18). Continuous measures were preferred over ICD-9/10 codes or binary thresholds, which may underestimate subclinical organ dysfunction in cohorts with healthy volunteer bias such as the UKBB(19).

### Z-Allele Dosage and Sensitivity Analyses

Using the same models, Z-allele dosage was modeled as zero (PIMM, PIMS, PISS), one (PIMZ, PISZ), or two (PIZZ) Z alleles. In sensitivity analyses, Z-allele status was modeled as binary (non-carriers vs. carriers), with results consistent with primary analyses (data not shown). An exploratory analysis assessed severe sarcopenia, requiring both low FFMI and low HGS(20).

### Causal Mediation Analysis and Mortality

Causal mediation analysis tested whether FFMI-defined sarcopenia mediated the association between additively modeled Z-allele dosage and all-cause mortality. Logistic regression was used to fit the mediator model (low FFMI ∼ Z-dosage) and the outcome model (all-cause mortality ∼ Z-dosage + low FFMI), adjusting for smoking status, age at UKBB assessment, sex, PC1-5, FEV1% predicted, and FIB4. The average causal mediation effect, average direct effect, total effect, and proportion mediated were estimated using the R mediation package, with mortality ascertained through 3/11/22.

Analyses were conducted using complete-case data. All tests were two-tailed and performed at a significance level of less than 0.05. Analyses were performed using R 4.5.2.

## Results

In 487,207 UKBB participants, genotype distribution was predominantly PI*MM (n=426,686), with smaller numbers of PI*MS (n=41,291), PI*SS (n=1,026), PI*MZ (n=17,170), PI*SZ (n=870), and PI*ZZ (n=164) (**Table 1**). Sex distribution was similar across groups (approximately 45-50% male; p=0.38). Race differed significantly (p<0.001), reflecting the expected predominance of White participants in deficient genotypes (≥99% White in PI*MS, PI*SS, PI*MZ, PI*SZ, and PI*ZZ compared with 93.4% in PI*MM) (21). Smoking status varied modestly across groups (p=0.02), though distributions were broadly similar; pack-years differed statistically (p=0.006), largely driven by lower mean exposure in PI*ZZ individuals (14.4 pack-years).

**Table 1 –.** Demographic characteristics of patients in the UK Biobank by AAT genotype.

|  | MM | MS | SS | MZ | SZ | ZZ | p value<sup>a</sup> |
| --- | --- | --- | --- | --- | --- | --- | --- |
| **n** | 426686 | 41291 | 1026 | 17170 | 870 | 164 |  |
| **Sex = Male (%)** | 195371 (45.8) | 18927 (45.8) | 476 (46.4) | 7794 (45.4) | 394 (45.3) | 82 (50.0) | 0.38 |
| **Body mass index (BMI) (mean (SD))** | 27.43 (4.79) | 27.41 (4.81) | 27.24 (4.64) | 27.27 (4.71)# | 27.02 (4.61)& | 26.81 (4.67) | <0.001 |
| **Age when attended assessment center (mean (SD))** | 56.51 (8.10) | 56.72 (8.03) | 56.38 (8.16) | 56.90 (8.08)# | 56.55 (7.81) | 56.26 (7.89) | <0.001 |
| **Ethnicity group (%)** |  |  |  |  |  |  | <0.001 |
| **White** | 398749 (93.4) | 40810 (98.8) | 1019 (99.3) | 17000 (99.0) | 865 (99.4) | 164 (100.0) |  |
| **Black** | 7547 (1.8) | 49 (0.1) | <10 | 19 (0.1) | <10 | <10 |  |
| **Asian** | 9351 (2.2) | 14 (0.0) | <10 | <10 | <10 | <10 |  |
| **Other** | 2588 (0.6) | 149 (0.4) | <10 | 55 (0.3) | <10 | <10 |  |
| **Missing** | 8451 (2.0) | 269 (0.7) | 6 (0.6) | 89 (0.5) | <10 | <10 |  |
| **Smoking status (%)<sup>w</sup>** |  |  |  |  |  |  | 0.02 |
| **Never** | 171039 (40.1) | 16091 (39.0) | 419 (40.8) | 6754 (39.3) | 317 (36.4) | 71 (43.3) |  |
| **Former** | 220018 (51.6) | 21905 (53.1) | 527 (51.4) | 9152 (53.3) | 483 (55.5) | 91 (55.5) |  |
| **Current** | 33417 (7.8) | 3135 (7.6) | 78 (7.6) | 1202 (7.0) | 66 (7.6) | <10 |  |
| **Missing** | 2212 (0.5) | 160 (0.4) | 2 (0.0) | 62 (0.4) | 4 (0.0) | 1 (0.0) |  |
| **Maternal smoking history** | 107273 (25.1) | 10809 (26.2) | 271 (26.4) | 4321 (25.2) | 226 (26.0) | 43 (26.2) | <0.001 |
| **Pack-years of smoking (mean (SD))<sup>x</sup>** | 23.32 (18.74) | 23.62 (19.02) | 25.52 (19.73) | 23.29 (19.36) | 22.58 (18.59) | 14.36 (8.87)\$ | 0.006 |
| **Subjects with pack-year data (%)** | 128616 (30.1) | 12799 (31.0) | 308 (30.1) | 5206 (30.3) | 262 (30.1) | 38 (23.2) |  |
| **Alcohol status (%)** |  |  |  |  |  |  | 0.010 |
| **Never** | 19595 (4.6) | 1368 (3.3) | 36 (3.5) | 617 (3.6) | 29 (3.3) | <10 |  |
| **Former** | 15361 (3.6) | 1454 (3.5) | 39 (3.8) | 600 (3.5) | 37 (4.3) | <10 |  |
| **Current** | 390703 (91.6) | 38420 (93.1) | 950 (92.6) | 15942 (92.8) | 804 (92.4) | 150 (91.5) |  |
| **Missing** | 1027 (0.24) | 49 (0.12) | 1 (0.1) | 11 (0.06) | 0 (0) | 0 (0) |  |
| **Total weekly alcohol units (mean (SD))** | 7.58 (8.70) | 7.48 (8.40) | 6.96 (7.88) | 7.18 (8.56)# | 7.43 (8.34) | 7.11 (7.37) | 0.074 |
| **Subjects with alcohol unit data (%)** | 98342 (23.1) | 9685 (23.5) | 239 (23.3) | 3967 (23.1) | 205 (23.6) | 54 (32.9) |  |
| **Comorbidities** |  |  |  |  |  |  |  |
| **Cancer\* (%)** | 32373 (7.6) | 3213 (7.8) | 80 (7.8) | 1330 (7.8) | 77 (8.9) | 13 (7.9) | 0.484 |
| **Allergic rhinitis\* (%)** | 23469 (5.5) | 2470 (6.0) | 55 (5.4) | 1024 (6.0) | 46 (5.3) | 11 (6.7) | 0.577 |
| **Asthma\* (%)** | 17872 (4.2) | 1861 (4.5) | 43 (4.2) | 777 (4.5) | 31 (3.6) | 11 (6.7) | 0.180 |
| **Alpha-1 antitrypsin deficiency (AATD)\* known diagnosis(%)** | 31 (0.1) | <10 | <10 | 39 (0.2)# | <10 | 16 (9.8)\$ | NA |
| **Bronchiectasis\* (%)** | 1082 (0.3) | 112 (0.3) | <10 | 54 (0.3) | <10 | <10 | NA |
| **Fractured bones in last 5 yrs (%)** | 40169 (9.4) | 3867 (9.4) | 104 (10.1) | 1728 (10.1)# | 92 (10.6) | 22 (13.4)\$ | 0.021 |
| **Liver disease (%)** | 1430 (0.3) | 131 (0.3) | <10 | 68 (0.4) | <10 | <10 | NA |
| **Liver cancer (%)** | 579 (0.1) | 57 (0.1) | <10 | 30 (0.2) | <10 | <10 | NA |
| **COPD defined by FEV1/FVC ratio <0.70** | 47504 (11.1) | 4898 (11.9) | 119 (11.6) | 2128 (12.4) | 123 (14.1)& | 49 (30.0)\$ | <0.001 |
| **COPD (composite definition)** | 49836 (11.7) | 5127 (12.4) | 129 (14.8)# | 2218 (12.9) | 124 (14.3)& | 56 (34.1)\$ | <0.001 |
| **Medications** |  |  |  |  |  |  |  |
| **Oral corticosteroids** | 4157 (1.0) | 424 (1.0) | <10 | 208 (1.2)# | 14 (1.6)& | <10 | NA |
| **Inhaled corticosteroids** | 11447 (2.7) | 1106 (2.7) | 33 (3.2) | 443 (2.6) | 26 (3.0) | <10 | NA |
| **Statin usage** | 72731 (17.0) | 7177 (17.4) | 167 (16.3) | 2778 (16.2)# | 171 (19.7)& | 15 (9.1)\$ | <0.001 |
| **Outcomes** |  |  |  |  |  |  |  |
| **Mortality** | 29386 (6.89) | 2852 (6.91) | 75 (7.31) | 1276 (7.43) | 71 (8.16) | 30 (18.3)\$ | <0.001 |

Regarding comorbidities, COPD prevalence (by FEV1/FVC <0.70) was higher in PI*ZZ versus PI*MM individuals (30.0% vs. 11.1%; p<0.05). A known diagnosis of liver disease or liver cancer was not significantly different among PI*MM vs. PI*ZZ, which may have been due to low overall counts. Fracture prevalence was modestly higher among PI*ZZ than PI*MM individuals (13.4% vs. 9.4%, p<0.05). Mortality over the time-frame analyzed (2006-2022) was also higher among PI*ZZ than PI*MM individuals (18.3% vs. 6.9%, p<0.05).

Overall, the most clinically meaningful findings were the increased prevalence of comorbidities and mortality among PI*ZZ individuals.

Across AAT genotypes, spirometric measures were largely similar among PI*MM, PI*MS, PI*SS, PI*MZ, and PI*SZ individuals, with mean FEV1% predicted approximately 95-96% and FEV1/FVC ratios approximately 0.76 (**Table 2**). However, PI*ZZ individuals demonstrated lower lung function (FEV1% predicted 88.6%, FEV1/FVC 0.71; p<0.05 compared to PI*MM). Respiratory symptoms were also more prevalent in PI*ZZ individuals compared to PI*MM. Laboratory parameters (CRP, liver function parameters) showed significant differences between PI*ZZ vs. PI*MM individuals. APRI, a non-invasive measure of hepatic fibrosis based on AST to platelet ratio(22), was significantly higher among PI*ZZ vs. PI*MM individuals (p<0.05). FIB4, a predictor of liver fibrosis risk, did not differ significantly as a continuous measure but was higher as a binary definition of liver fibrosis risk(18).

**Table 2 –.** Spirometry, symptoms, and labs for AAT genotypes.

|  | MM | MS | SS | MZ | SZ | ZZ | p value<sup>a</sup> |
| --- | --- | --- | --- | --- | --- | --- | --- |
| **n** | 426686 | 41291 | 1026 | 17170 | 870 | 164 |  |
| **Spirometry** |  |  |  |  |  |  |  |
| **FEV1% percent predicted** | 95.61 (16.87) | 95.74 (16.92) | 95.67 (15.74) | 96.11 (17.30)# | 95.58 (17.30) | 88.61 (24.66)\$ | 0.035 |
| **FEV1/FVC ratio (mean (SD))** | 0.76 (0.07) | 0.76 (0.07) | 0.76 (0.07) | 0.76 (0.07) | 0.76 (0.08) | 0.71 (0.13)\$ | <0.001 |
| **FEV1 (liters)** | 2.81 (0.80) | 2.85 (0.80) | 2.87 (0.76) | 2.89 (0.79)# | 2.92 (0.78) | 2.73 (1.00)\$ | <0.001 |
| **Symptoms** |  |  |  |  |  |  |  |
| **Shortness of breath on level ground (%)** | 16516 (11.2) | 1470 (10.7) | 30 (8.3) | 596 (10.3) | 29 (9.7) | 23 (31.0)\$ | <0.001 |
| **Subjects with shortness of breath data (%)** | 146714 (34.3) | 13723 (33.2) | 360 (35.1) | 5749 (33.5) | 296 (34.0) | 74 (45) |  |
| **Cough on most days (%)** | 13986 (13.6) | 1481 (14.1) | 23 (9.7) | 585 (13.1) | 30 (13.0) | 19 (40.4)\$ | <0.001 |
| **Subjects with cough data (%)** | 103123 (24.2) | 10496 (25.4) | 237 (23.1) | 4463 (26.0) | 231 (26.6) | 47 (28.7) |  |
| **Bring up sputum on most days (%)** | 8893 (8.6) | 946 (9.0) | 13 (5.5) | 353 (7.9) | 20 (8.7) | 15 (31.9)\$ | <0.001 |
| **Subjects with sputum data (%)** | 103123 (24.2) | 10496 (25.4) | 237 (23.1) | 4463 (26.0) | 231 (26.6) | 47 (28.7) |  |
| **Labs** |  |  |  |  |  |  |  |
| **C reactive protein (mean (SD))\*** | 2.61 (4.35) | 2.56 (4.37) | 2.32 (3.54) | 2.44 (4.43)# | 2.15 (3.21) | 2.80 (6.90)\$ | <0.001 |
| **Vitamin D (mean (SD))** | 48.51 (21.12) | 49.46 (21.11) | 49.26 (21.09) | 49.57 (20.79)# | 49.79 (20.89)& | 47.54 (19.12)\$ | <0.001 |
| **Albumin (mean (SD))** | 45.16 (2.62) | 45.45 (2.61) | 45.85 (2.57) | 45.89 (2.62)# | 46.13 (2.57)& | 45.65 (2.72)\$ | <0.001 |
| **AST (mean (SD))\*** | 24.87 (11.95) | 24.91 (11.26) | 25.52 (11.57) | 26.04 (10.93)# | 25.65 (11.38) | 29.49 (13.84)\$ | <0.001 |
| **ALT (mean (SD))\*** | 22.36 (14.74) | 22.68 (14.55) | 23.44 (14.82) | 24.00 (13.76)# | 23.86 (14.69)& | 24.45 (13.97)\$ | <0.001 |
| **GGT (mean (SD))** | 35.60 (41.95) | 35.88 (42.70) | 35.69 (38.49) | 36.52 (33.99) | 36.39 (36.16) | 40.24 (51.06) | 0.478 |
| **Bilirubin (mean (SD))\*** | 8.65 (4.75) | 8.71 (4.78) | 8.79 (4.79) | 8.92 (4.98)# | 8.86 (4.85) | 8.81 (4.71) | <0.001 |
| **Sodium (mean (SD))** | 1.33 (0.58) | 1.34 (0.58) | 1.33 (0.58) | 1.38 (0.56)# | 1.34 (0.57) | 1.30 (0.65) | 0.094 |
| **Prothrombin time (mean (SD))** | 13.08 (2.53) | 13.09 (2.43) | 13.12 (2.45) | 13.06 (2.49) | 13.08 (2.43) | 13.21 (2.01) | 0.499 |
| **Platelets (mean (SD))\*** | 252.77 (59.98) | 255.16 (59.85) | 255.07 (61.72) | 254.27 (59.71)# | 255.31 (61.12) | 238.18 (64.62)\$ | <0.001 |
| **Total cholesterol mmol/L (mean(SD))** | 5.69 (1.15) | 5.74 (1.14) | 5.72 (1.13) | 5.74 (1.15)# | 5.72 (1.15) | 5.91 (1.07)\$ | <0.001 |
| **LDL mmol/L (mean (SD))** | 3.55 (0.89) | 3.59 (0.87) | 3.58 (0.86) | 3.61 (0.88)# | 3.57 (0.87) | 3.69 (0.81)\$ | <0.001 |
| **HDL mmol/L (mean (SD))** | 1.45 (0.38) | 1.45 (0.38) | 1.45 (0.39) | 1.43 (0.38)# | 1.47 (0.38) | 1.58 (0.49)\$ | <0.001 |
| **Triglycerides mmol/L (mean (SD))** | 1.75 (1.03) | 1.77 (1.03) | 1.73 (1.01) | 1.76 (1.02) | 1.70 (1.00) | 1.48 (0.76)\$ | <0.001 |
| **Apolipoprotein A mg/dL (mean (SD))** | 72.3 (18.6) | 72.1 (18.4) | 72.2 (19.6) | 71.5 (15.8) | 72.0 (15.3) | 71.3 (13.3) | <0.001 |
| **Apolipoprotein B g/L (mean (SD))** | 1.03 (0.24) | 1.04 (0.24) | 1.03 (0.24) | 1.05 (0.24) | 1.02 (0.24) | 1.05 (0.22) | <0.001 |
| **Noninvasive liver indices** |  |  |  |  |  |  |  |
| **APRI** | 1.34 (1.97) | 1.32 (0.81) | 1.34 (0.70) | 1.34 (0.66) | 1.31 (0.61) | 1.63 (0.77)\$ | 0.126 |
| **AST/ALT ratio\*** | 1.26 (0.43) | 1.24 (0.43) | 1.22 (0.42) | 1.22 (0.46)# | 1.21 (0.41)& | 1.32 (0.41) | <0.001 |
| **FIB-4** | 0.27 (0.50) | 0.26 (0.23) | 0.27 (0.19) | 0.28 (0.19) | 0.27 (0.16) | 0.34 (0.20) | 0.167 |
| **High FIB-4** | 8964 (2.1) | 785 (1.9) | 21 (2.0) | 367 (2.1) | 17 (2.0) | 13 (7.9)\$ | <0.001 |
| **High APRI** | 845 (0.2) | 54 (0.1) | <10 | 33 (0.2) | <10 | <10 | NA |
| **High APRI and FIB4** | 789 (0.2) | 50 (0.1) | <10 | 32 (0.2) | <10 | <10 | NA |

<sup>a</sup> p-value represents A-NOVA for continuous variables and chisquare for categorical variables.
Non-invasive markers of hepatic fibrosis were calculated using baseline laboratory values. The AST-to-Platelet Ratio Index (APRI) was computed as (AST/ULN)/platelet count × 100, using an upper limit of normal (ULN) for AST of 40 U/L and platelet count expressed in 10<sup>9</sup>/L. The Fibrosis-4 index (FIB-4) was calculated as (age × AST)/(platelet count × √ALT), incorporating age (years), AST (U/L), ALT (U/L), and platelet count. The AST/ALT ratio was calculated as AST divided by ALT. High FIB-4 was defined as >2.67 as defined in the literature.

In terms of body composition and physical function, several parameters differed across *SERPINA1* genotypes (**Table 3**). Slow walking pace showed the strongest genotype association, with the highest prevalence observed in PI*ZZ compared to PI*MM (12.8% versus 8.1%, p<0.05), consistent with greater functional impairment in severe AATD. Measures of muscle mass also differed by genotype. Compared with PI*MM individuals, PI*MZ individuals had slightly higher whole-body fat mass but lower whole-body fat-free mass, FFMI, and ASMI compared to PI*MM (all p<0.05). Similarly, both PI*SZ and PI*ZZ individuals demonstrated lower FFMI and ASMI and a higher prevalence of FFMI-defined sarcopenia compared with PI*MM (all p<0.05; **Figures 1-2**). HGS was broadly similar across genotypes, ranging from 31.8-32.6 kg, although PI*MZ individuals had slightly HGS (∼0.35 kg) than PI*MM (p<0.05). Physical activity measures were similar across *SERPINA1* genotypes. Weight loss in the prior year differed modestly across genotypes (p<0.001) but did not demonstrate a clear Z-allele gradient.

**Figure 2.**
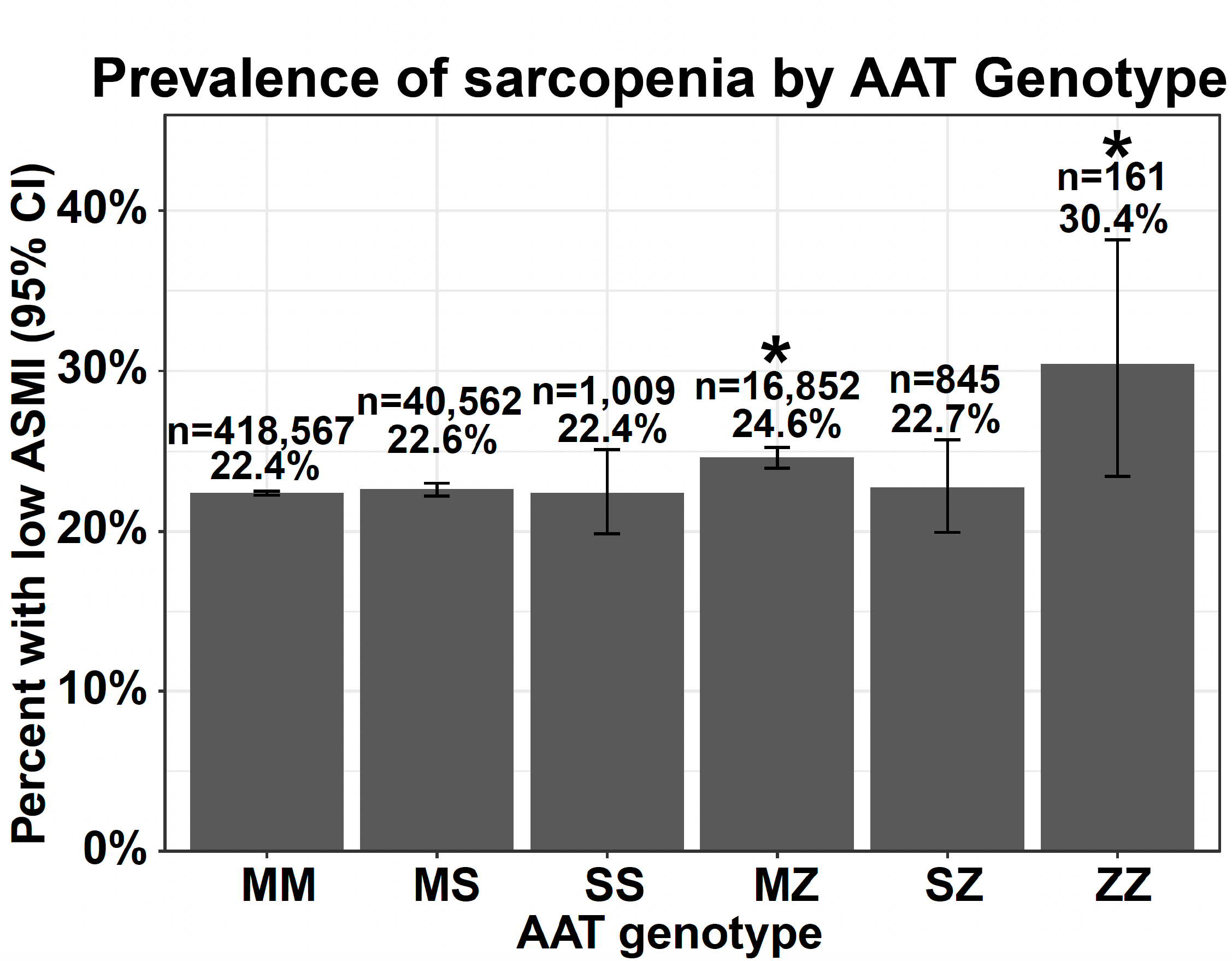
Prevalence of ASMI-defined sarcopenia across AAT genotypes. ASMI-defined sarcopenia prevalence demonstrated a genotype-dependent pattern, with progressively higher proportions observed among Z allele carriers and the highest prevalence in ZZ individuals (30.4%). Error bars represent 95% confidence intervals. Sample sizes are indicated above each bar. * indicates statistically significant prevalence by Z test of proportions compared to PI*MM.

**Table 3 –.**
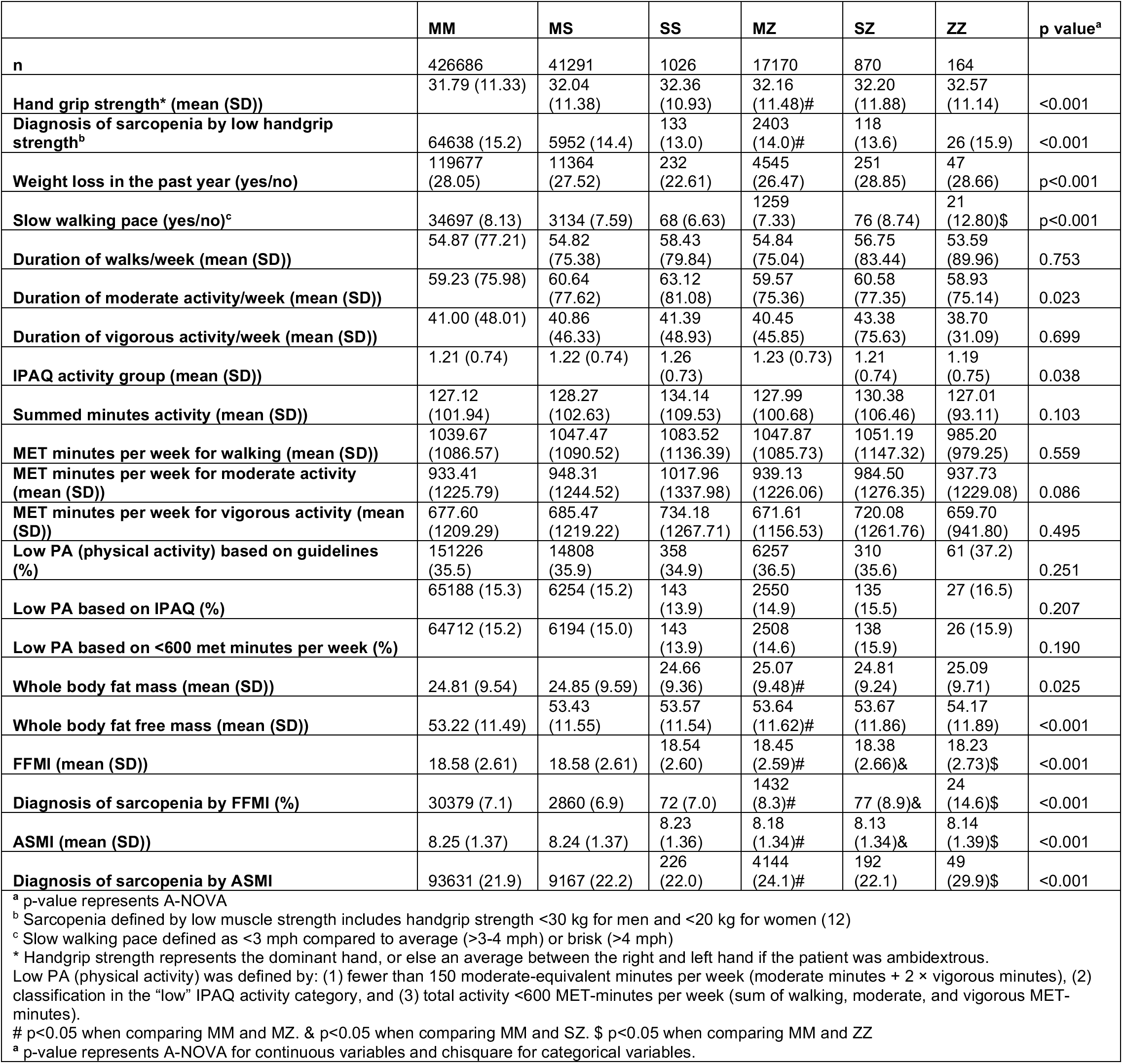
Body composition and physical activity parameters by AAT genotype.

|  | MM | MS | SS | MZ | SZ | ZZ | p value<sup>a</sup> |
| --- | --- | --- | --- | --- | --- | --- | --- |
| **n** | 426686 | 41291 | 1026 | 17170 | 870 | 164 |  |
| **Hand grip strength\* (mean (SD))** | 31.79 (11.33) | 32.04 (11.38) | 32.36 (10.93) | 32.16 (11.48)# | 32.20 (11.88) | 32.57 (11.14) | <0.001 |
| **Diagnosis of sarcopenia by low handgrip strength<sup>b</sup>** | 64638 (15.2) | 5952 (14.4) | 133 (13.0) | 2403 (14.0)# | 118 (13.6) | 26 (15.9) | <0.001 |
| **Weight loss in the past year (yes/no)** | 119677 (28.05) | 11364 (27.52) | 232 (22.61) | 4545 (26.47) | 251 (28.85) | 47 (28.66) | p<0.001 |
| **Slow walking pace (yes/no)<sup>c</sup>** | 34697 (8.13) | 3134 (7.59) | 68 (6.63) | 1259 (7.33) | 76 (8.74) | 21 (12.80)\$ | p<0.001 |
| **Duration of walks/week (mean (SD))** | 54.87 (77.21) | 54.82 (75.38) | 58.43 (79.84) | 54.84 (75.04) | 56.75 (83.44) | 53.59 (89.96) | 0.753 |
| **Duration of moderate activity/week (mean (SD))** | 59.23 (75.98) | 60.64 (77.62) | 63.12 (81.08) | 59.57 (75.36) | 60.58 (77.35) | 58.93 (75.14) | 0.023 |
| **Duration of vigorous activity/week (mean (SD))** | 41.00 (48.01) | 40.86 (46.33) | 41.39 (48.93) | 40.45 (45.85) | 43.38 (75.63) | 38.70 (31.09) | 0.699 |
| **IPAQ activity group (mean (SD))** | 1.21 (0.74) | 1.22 (0.74) | 1.26 (0.73) | 1.23 (0.73) | 1.21 (0.74) | 1.19 (0.75) | 0.038 |
| **Summed minutes activity (mean (SD))** | 127.12 (101.94) | 128.27 (102.63) | 134.14 (109.53) | 127.99 (100.68) | 130.38 (106.46) | 127.01 (93.11) | 0.103 |
| **MET minutes per week for walking (mean (SD))** | 1039.67 (1086.57) | 1047.47 (1090.52) | 1083.52 (1136.39) | 1047.87 (1085.73) | 1051.19 (1147.32) | 985.20 (979.25) | 0.559 |
| **MET minutes per week for moderate activity (mean (SD))** | 933.41 (1225.79) | 948.31 (1244.52) | 1017.96 (1337.98) | 939.13 (1226.06) | 984.50 (1276.35) | 937.73 (1229.08) | 0.086 |
| **MET minutes per week for vigorous activity (mean (SD))** | 677.60 (1209.29) | 685.47 (1219.22) | 734.18 (1267.71) | 671.61 (1156.53) | 720.08 (1261.76) | 659.70 (941.80) | 0.495 |
| **Low PA (physical activity) based on guidelines (%)** | 151226 (35.5) | 14808 (35.9) | 358 (34.9) | 6257 (36.5) | 310 (35.6) | 61 (37.2) | 0.251 |
| **Low PA based on IPAQ (%)** | 65188 (15.3) | 6254 (15.2) | 143 (13.9) | 2550 (14.9) | 135 (15.5) | 27 (16.5) | 0.207 |
| **Low PA based on <600 met minutes per week (%)** | 64712 (15.2) | 6194 (15.0) | 143 (13.9) | 2508 (14.6) | 138 (15.9) | 26 (15.9) | 0.190 |
| **Whole body fat mass (mean (SD))** | 24.81 (9.54) | 24.85 (9.59) | 24.66 (9.36) | 25.07 (9.48)# | 24.81 (9.24) | 25.09 (9.71) | 0.025 |
| **Whole body fat free mass (mean (SD))** | 53.22 (11.49) | 53.43 (11.55) | 53.57 (11.54) | 53.64 (11.62)# | 53.67 (11.86) | 54.17 (11.89) | <0.001 |
| **FFMI (mean (SD))** | 18.58 (2.61) | 18.58 (2.61) | 18.54 (2.60) | 18.45 (2.59)# | 18.38 (2.66)& | 18.23 (2.73)\$ | <0.001 |
| **Diagnosis of sarcopenia by FFMI (%)** | 30379 (7.1) | 2860 (6.9) | 72 (7.0) | 1432 (8.3)# | 77 (8.9)& | 24 (14.6)\$ | <0.001 |
| **ASMI (mean (SD))** | 8.25 (1.37) | 8.24 (1.37) | 8.23 (1.36) | 8.18 (1.34)# | 8.13 (1.34)& | 8.14 (1.39)\$ | <0.001 |
| **Diagnosis of sarcopenia by ASMI** | 93631 (21.9) | 9167 (22.2) | 226 (22.0) | 4144 (24.1)# | 192 (22.1) | 49 (29.9)\$ | <0.001 |

<sup>a</sup> p-value represents A-NOVA
<sup>b</sup> Sarcopenia defined by low muscle strength includes handgrip strength <30 kg for men and <20 kg for women (12)
<sup>c</sup> Slow walking pace defined as <3 mph compared to average (>3-4 mph) or brisk (>4 mph)
\* Handgrip strength represents the dominant hand, or else an average between the right and left hand if the patient was ambidextrous.
Low PA (physical activity) was defined by: (1) fewer than 150 moderate-equivalent minutes per week (moderate minutes + 2 × vigorous minutes), (2) classification in the “low” IPAQ activity category, and (3) total activity <600 MET-minutes per week (sum of walking, moderate, and vigorous MET-minutes).

Unbiased PheWAS of our dataset was then performed to determine whether the AAT Z allele (genotype) was associated with clinical phenotypes in the UKBB (**Figure 3**, **Supp Table 1**). Characteristic features associated with the AAT Z allele included albumin, globulin, AAT diagnosis, liver parameters (AST, ALT, GGT) and lung parameters (FVC, emphysema diagnosis). However, a number of measures related to body composition (muscle, bone, and fat) were also significantly associated with the Z allele, prompting further analysis with univariate and multivariate regression models to determine whether Z-associated genotypes were associated with lower muscle mass and binary-definitions of sarcopenia.

**Figure 3.**
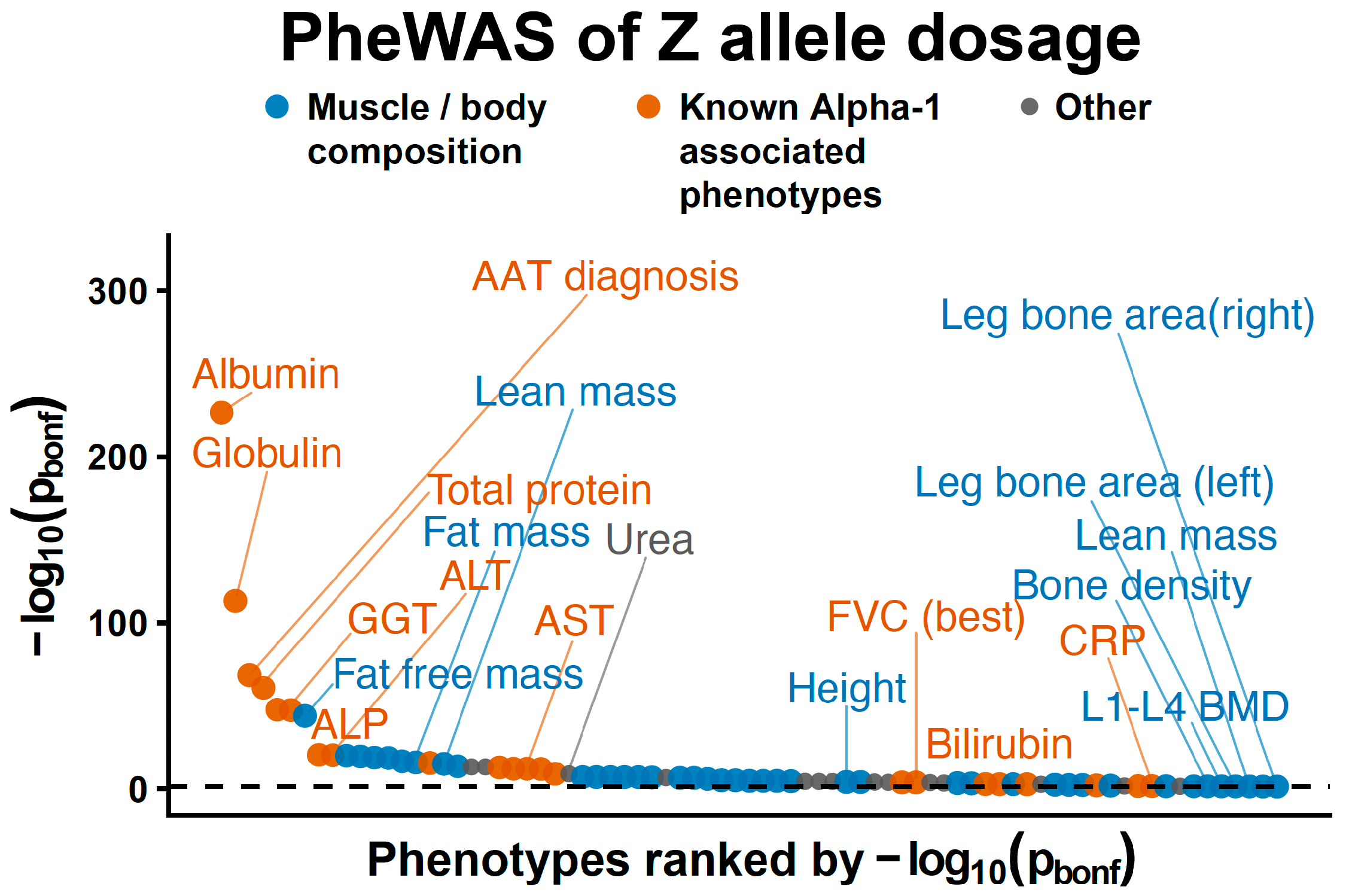
PheWAS of Z allele dosage. Phenome-wide association analysis of Z allele dosage across UK Biobank phenotypes ranked by −log10(p). The y-axis displays the negative base-10 logarithm of the p-value (−log₁₀[p]) for the association between Z allele and the phenotype. Higher values indicate stronger statistical evidence of association. The dashed horizontal line corresponds to p < 0.05 (−log₁₀ > 1.3) with Bonferroni adjustment. The x-axis represents phenotypes ranked by statistical significance (descending −log₁₀[p bonferroni]), such that the most statistically significant phenotype is positioned first, followed sequentially by less significant phenotypes. Muscle/bone/body composition and known alpha-1 antitrypsin–associated phenotypes are highlighted, and significant associations are labeled. The dashed horizontal line represents the phenome-wide significance threshold.

Compared with PI*MM, PI*MZ, PI*SZ, and PI*ZZ genotypes were associated with lower FFMI and ASMI after adjustment for core covariates, lung function (FEV1% predicted), and liver function (FIB4) (**Figure 4A**, **Supp Table 2 A**). When using binary definitions of sarcopenia and logistic regression models, PI*MZ heterozygotes demonstrated increased odds of sarcopenia defined by both FFMI and ASMI. These associations persisted after full adjustment for core covariates, lung function, and liver function. PI*ZZ individuals demonstrated the strongest association with FFMI-defined sarcopenia, with approximately twofold higher odds that remained significant after full adjustment (adjusted OR 2.02, 95% CI 1.16–3.32). In contrast, associations with the PI*SZ genotype were modest and were only significant for FFMI-defined sarcopenia in fully adjusted models but not for ASMI-defined sarcopenia in fully adjusted models, while PI*MS and PI*SS genotypes were not significantly associated with binary-defined sarcopenia across all models. Overall, the most consistent and robust associations with sarcopenia were observed with the PI*MZ and PI*ZZ genotypes (**Figure 4B**, **Supp Table 2 B**).

**Figure 4.**
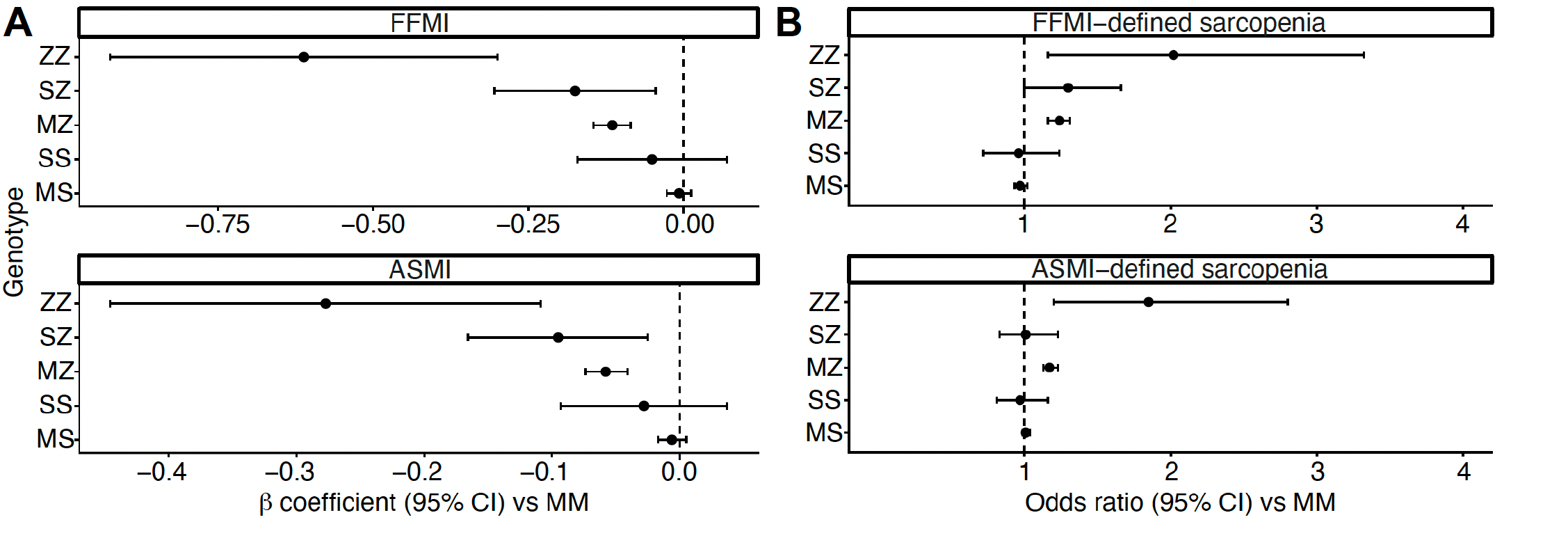
Z-related AAT genotypes are associated with reduced lean mass and increased odds of sarcopenia. Left panel: AAT genotypes PI*MZ, PI*SZ, and PI*ZZ were associated with lower FFMI and ASMI in fully adjusted linear regression models, with associations persisting after adjustment for lung function (FEV1% predicted) and liver function (FIB4). Right panel: AAT genotypes PI*MZ and PI*ZZ were associated with increased odds of FFMI- and ASMI-defined sarcopenia in fully adjusted models. Points represent β coefficients for FFMI or ASMI (as a continuous variable) or odds ratios of binary definitions for FFMI or ASMI sarcopenia with 95% confidence intervals. Dashed vertical lines indicate the null value (β = 0, OR = 1). Estimates whose 95% confidence intervals cross this line are not statistically significant. Adjusted model = adjusted for core covariates (sex, smoking status, age at assessment, PC1-5). *Adjusted model = adjusted for core covariates, lung function (FEV1% predicted), and liver function (FIB-4).

We then performed a Z allele dosage analysis comparing genotypes with zero Z alleles (PI*MM, PI*MS, PI*SS), one Z allele (PI*MZ, PI*SZ), and two Z alleles (PI*ZZ). Each Z-allele was significantly associated with reduced FFMI and ASMI in fully adjusted linear models incorporating lung function (FEV1% predicted) and liver function (FIB4). Compared with zero Z alleles, one Z allele was associated with lower muscle mass, while two Z alleles was associated with a greater decrease in muscle mass. Consistent with these findings, one Z allele was associated with higher odds of sarcopenia, while two Z alleles were associated with higher odds of sarcopenia compared to PI*MM. Overall, these results support an association between the Z allele and reduced muscle mass that persisted after adjustment for pulmonary and hepatic function (**Figure 5A-B**, **Supp Table 3A-B**).

**Figure 5.**
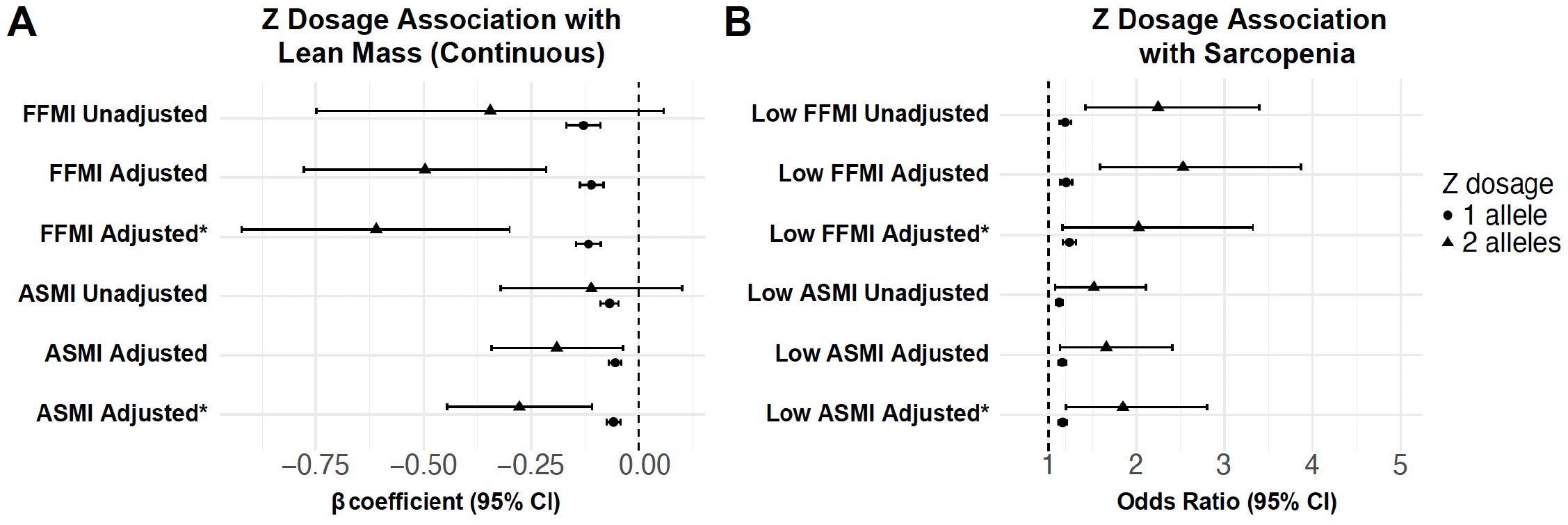
The Z Allele Is associated with reduced lean mass and increased odds of sarcopenia. Left panel: Genotypes with one Z allele (PI*MZ, PI*SZ) and two Z alleles (PI*ZZ) were associated with lower FFMI and ASMI in unadjusted and adjusted linear regression models, with associations persisting after adjustment for lung function (FEV1% predicted) and liver function (FIB4). Right panel: Genotypes with one Z allele or two Z alleles were associated with increased odds of FFMI- and ASMI-defined sarcopenia in fully adjusted models. Points represent β coefficients or odds ratios with 95% confidence intervals; dashed vertical lines indicate the null value (β = 0, OR = 1). Adjusted model = adjusted for core covariates (sex, smoking status, age at assessment, PC1-5). *Adjusted model = adjusted for core covariates, lung function (FEV1% predicted), and liver function (FIB-4).

When using a definition of severe sarcopenia requiring both low FFMI and low HGS(20), the PI*ZZ genotype was associated with increased odds of severe sarcopenia in unadjusted and core covariate-adjusted models, although this association was attenuated and no longer statistically significant after adjustment for lung and liver function. PI*MZ genotype showed a modest increase in odds in unadjusted and adjusted models, which did not persist in models adjusted for lung and liver function. Z-allele dosage was associated with increased odds of sarcopenia in unadjusted and core-adjusted models, particularly among individuals with two Z alleles. These associations were attenuated and no longer significant after adjustment for lung and liver function (**Supp Table 4-5**).

**Table 4 –.** Mediation Analysis Evaluating FFMI-defined Sarcopenia as a Mediator of the Z-Allele and All-Cause Mortality.

| Model | Association | OR (95% CI) | p value |
| --- | --- | --- | --- |
| **Total effect** | Z dosage → mortality | **1.086 (1.017-1.158)** | **0.013** |
| **Direct effect** | Z dosage → mortality (adjusted for FFMI-defined sarcopenia) | **1.083 (1.014-1.157)** | **0.017** |
| **Mediator model** | Z dosage → FFMI-defined sarcopenia | **1.249 (1.178-1.322)** | **<0.001** |
| **Outcome model** | FFMI-defined sarcopenia → mortality | **1.075 (1.022-1.13)** | **0.005** |
| Model represents a multivariate model adjusted for core covariates which include: age when attended assessment center, sex, smoking status (current / former / never), UK Biobank assessment center, PCs 1-5, FEV1% predicted, and FIB4. Ethnicity group removed because all ethnicity was White for certain AAT genotypes. |  |  |  |

Next, the association between the *SERPINA1* Z-allele and all-cause mortality was evaluated, and whether sarcopenia mediated this relationship. We determined that each Z allele was associated with increased mortality and higher odds of sarcopenia. Sarcopenia independently predicted mortality and modestly mediated the association between Z-allele dosage and mortality (∼2.6%; see **Table 4**).

## Discussion

In this population-based AATD analysis of the UKBB, PI*MZ and PI*ZZ genotypes were associated with reduced skeletal muscle mass and increased sarcopenia prevalence, independent of lung and liver function. Z-allele carriage was also associated with increased mortality, partially mediated by sarcopenia. These findings expand the phenotypic spectrum of AATD to include skeletal muscle.(7) (**Figure 6**).

**Figure 6.**
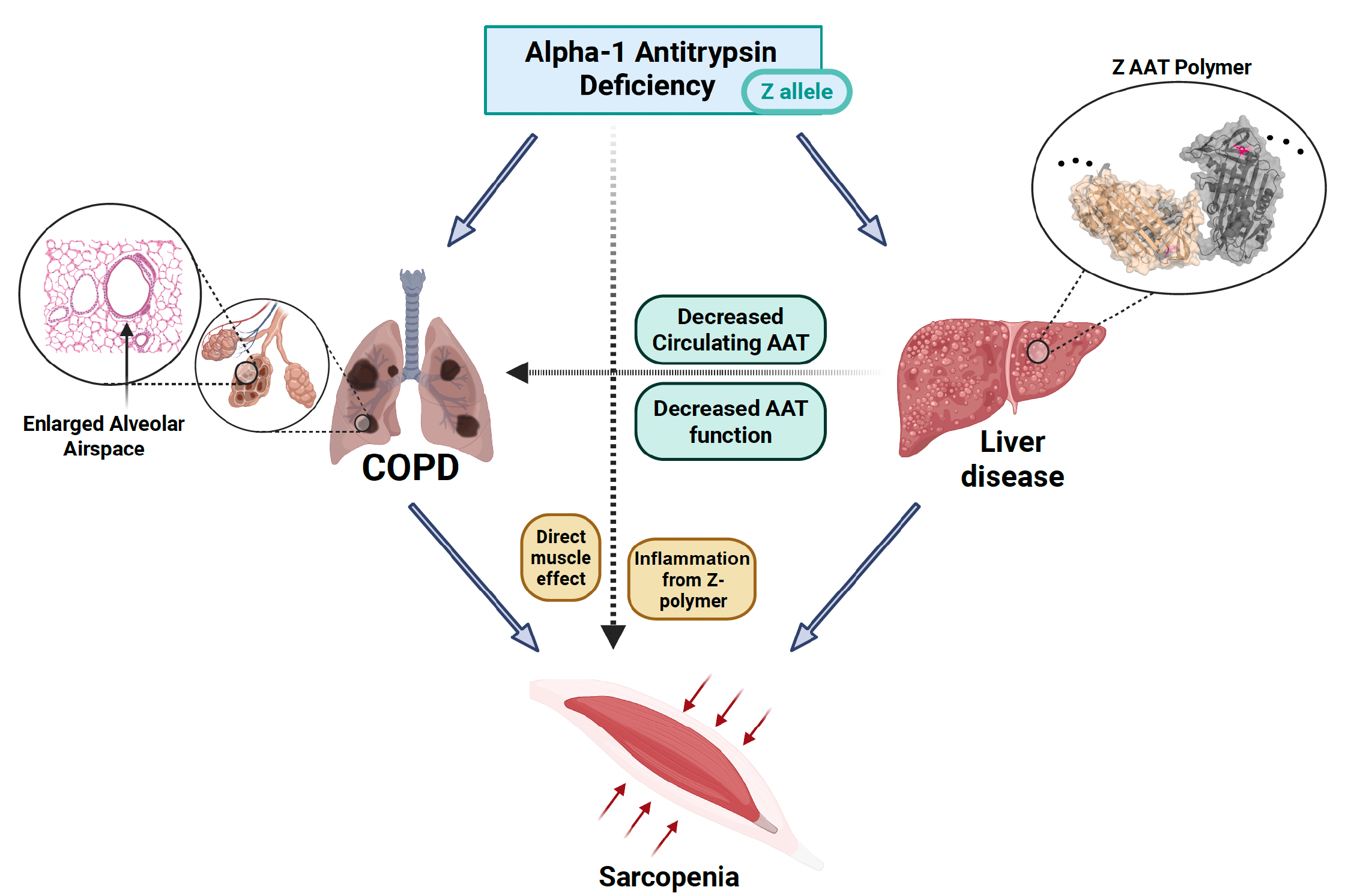
Proposed mechanisms linking alpha-1 antitrypsin deficiency (AATD) to sarcopenia. The *SERPINA1* Z allele leads to reduced circulating AAT levels and impaired AAT function, contributing to both lung and liver disease. In the lung, unopposed neutrophil-derived serine proteases promote alveolar destruction and emphysema. In the liver, accumulation of misfolded Z-AAT polymers cause hepatic injury and liver disease. Z-polymers are also secreted into the circulation and contribute to systemic inflammation. These processes may contribute to skeletal muscle loss through both indirect and direct pathways. Together, these mechanisms may converge to increase the risk of sarcopenia in AATD patients. Two units of Z AAT (in tan and grey) (PDB ID 3T1P, [ref: Yamasaki et al (2011) EMBO J]) are shown with Z mutation colored as before. Structural images were rendered with PyMOL v.2.6.2 [ref: The PyMOL Molecular Graphics System, Schrodinger LLC]. Additional images provided by BioRender.

Several potential mechanisms may explain the association between the Z allele and sarcopenia. Z polymers promote inflammation(23), which is a known driver of sarcopenia(24). Specifically, elevated CRP, IL-6, and TNF-α levels are consistently associated with lower skeletal muscle mass and strength(25). The inflammation caused by Z polymers may therefore promote sarcopenia through activation of NF-κB signaling or an impaired anabolic response to exercise and nutrition(26). Deficiency in AAT also causes unopposed activity of neutrophil-derived serine proteases (i.e. neutrophil elastase, proteinase 3, and cathepsin G), which may contribute to the observed association with sarcopenia(7). In other primary diseases of the muscle like muscular dystrophy, neutrophil elastase has been shown to impair myoblast survival, proliferation, and differentiation, contributing to loss of muscle regenerative capacity(27). Taken together, these mechanisms suggest that the Z allele may promote sarcopenia through both inflammatory and proteolytic pathways that impair muscle maintenance and regeneration.

The dose-dependent relationship between the Z-allele and sarcopenia risk support a gene-dosage effect consistent with AATD biology. The increase in sarcopenia risk across Z genotypes suggests an allele gradient where both loss-of-function (reduced circulating AAT) and gain-of-function (polymer accumulation) mechanisms may contribute to skeletal muscle pathology. Notably, the finding that PI*MZ heterozygotes demonstrated significantly lower FFMI and ASMI compared with PI*MM individuals is particularly relevant given the high population prevalence of this genotype (2–4% among individuals of European ancestry) (16). Although the magnitude of muscle loss at the individual level was modest, the population-attributable burden may be substantial. Importantly, genotypes without a Z-allele (e.g., PI*MS and PI*SS), which do not generate circulating Z-polymers or the same degree of systemic inflammation, were not consistently associated with sarcopenia. This suggests that the observed associations are specific to Z-allele biology rather than variations in the *SERPINA1* gene. Our findings align with emerging evidence that PI*MZ heterozygosity represents a distinct COPD endotype(28, 29, 30) and raises the possibility that it may similarly represent a sarcopenia endotype. These findings highlight the need for mechanistic studies to determine whether muscle impairment in those with a Z allele results from systemic inflammation due to circulating Z-polymers, direct toxic effects of Z-polymers to skeletal muscle, or unopposed neutrophil-derived serine protease activity due to AAT deficiency.

While our findings align with prior work reporting an association between AATD and muscle loss or dysfunction(20, 31, 32), the biologic mechanisms driving sarcopenia remain unclear. Others have demonstrated low muscle mass among patients with COPD due to AATD PI*ZZ compared with control PI*MM patients without COPD matched by age, sex, and smoking pack-year history. In addition, a study of AATD patients undergoing pulmonary rehabilitation (PR) demonstrated that PI*ZZ COPD patients achieved approximately 50% of the peak work rate observed in PI*MM COPD patients after three weeks of PR(31).

Skeletal muscle biopsies from these patients showed that PI*ZZ patients did not increase the proportion of Type I muscle fibers (which have greater oxidative capacity than Type II fibers(20)) following PR, whereas PI*MM patients increased their Type I muscle fiber proportion(32). These data suggest that individuals with PI*ZZ AATD are at increased risk for sarcopenia and exhibit impaired skeletal muscle functional response to PR. Our findings complement the recent work of Schrader et al.(33), who demonstrate that obesity and diabetes amplify hepatic risk in PI*ZZ individuals, whereas overweight status may be protective for pulmonary outcomes. By showing that the Z-allele independently associates with loss of fat-free mass, our study suggests that body composition phenotyping using measures such as FFMI and ASMI are essential to understanding organ-specific risks in AATD. We also noted that associations with severe sarcopenia were attenuated after adjustment, suggesting that functional impairment may be more strongly influenced by lung and liver disease than muscle mass alone.

While our analysis identified novel findings related to PI*Z genotype in a large, population-based cohort, several limitations of the study warrant comment. First, the UKBB is known to have a healthy volunteer bias, and the prevalence of severe AATD-related disease is likely underestimated compared to a clinically-diagnosed population. This bias may explain the small proportion of mortality that was mediated by sarcopenia from the Z allele and highlights the need for future studies in a clinically referred cohort with more severe disease. Second, body composition was assessed by bioelectrical impedance analysis rather than by gold standard measures such as dual-energy X-ray absorptiometry(20). Third, although we adjusted for FEV₁, we did not have imaging-based measures of emphysema and therefore could not account for its contribution in the UKBB. Given that emphysema is more negatively associated with muscle mass than airflow obstruction(34) and that emphysema demonstrates a dose-response with the Z-allele(29), incorporating quantitative emphysema measures will be an important future analysis. Finally, we acknowledge that lung and liver dysfunction may lie on the causal pathway between the *SERPINA1* Z-allele and sarcopenia. Therefore, adjustment for FEV₁ and FIB4 may attenuate the total effect of the Z-allele. Given the lack of data on circulating AAT levels and Z-polymer burden in the UKBB, future studies are needed to delineate the relative contributions of loss-of-function and gain-of-function pathways in sarcopenia due to AATD.

Despite these limitations, our study has important clinical implications. Although current guidelines recommend that all patients with COPD undergo AATD testing(35), only a small proportion of individuals in the UK Biobank had a documented diagnosis of AATD. Our findings that the Z-allele independently associates with sarcopenia and mortality further underscore the urgency of closing this diagnostic gap, as these associations can only inform clinical care if Z-alleles are identified(20). Our findings also suggest that interventions targeting sarcopenia, which include pulmonary rehabilitation, nutritional optimization with adequate protein intake, and resistance exercise may be particularly important in AATD populations. Given the high prevalence of sarcopenia observed among PI*ZZ individuals, these patients may represent a subgroup at heightened risk for functional decline. Emerging data suggesting that higher-dose augmentation therapy can reduce circulating inflammatory cytokines implicated in cachexia raises the possibility that higher augmentation dosage could influence muscle-related outcomes as well(36). Together, these findings suggest that muscle-related endpoints may warrant consideration in future studies of AAT-related disease and emerging therapies(37).

### Funded in part by

NIH K08 HL168348 (AA); Alpha 1 foundation grant

### Author contribution section

AA had full access to the data in the study and takes responsibility for the integrity of the data and the accuracy of the data analysis. Further details of author contributions are as follows: study concept and design, AA, JS, RB, JB, KS, SD, UH, JZ, PB; acquisition of data, AA, PB; statistical analysis, AA, LR, PB, JZ, VO; drafting of the first version of the manuscript, AA, KS, SD, UH, JZ, PB, MK, JD, LR, RW, VT, JB, JW, VO, JS, RB. All authors contributed to interpretation of data and critical revision of the manuscript for important intellectual content and agree to be accountable for all aspects of the work.

## Data Availability

Access to the underlying UK Biobank data is available to approved researchers through the UK Biobank Research Analysis Platform. This study was conducted under UK Biobank application number 61825.

## Acknowledgement.

Access to the UK Biobank dataset was obtained through application number 61825.

---

**Supplementary Figure 1.**
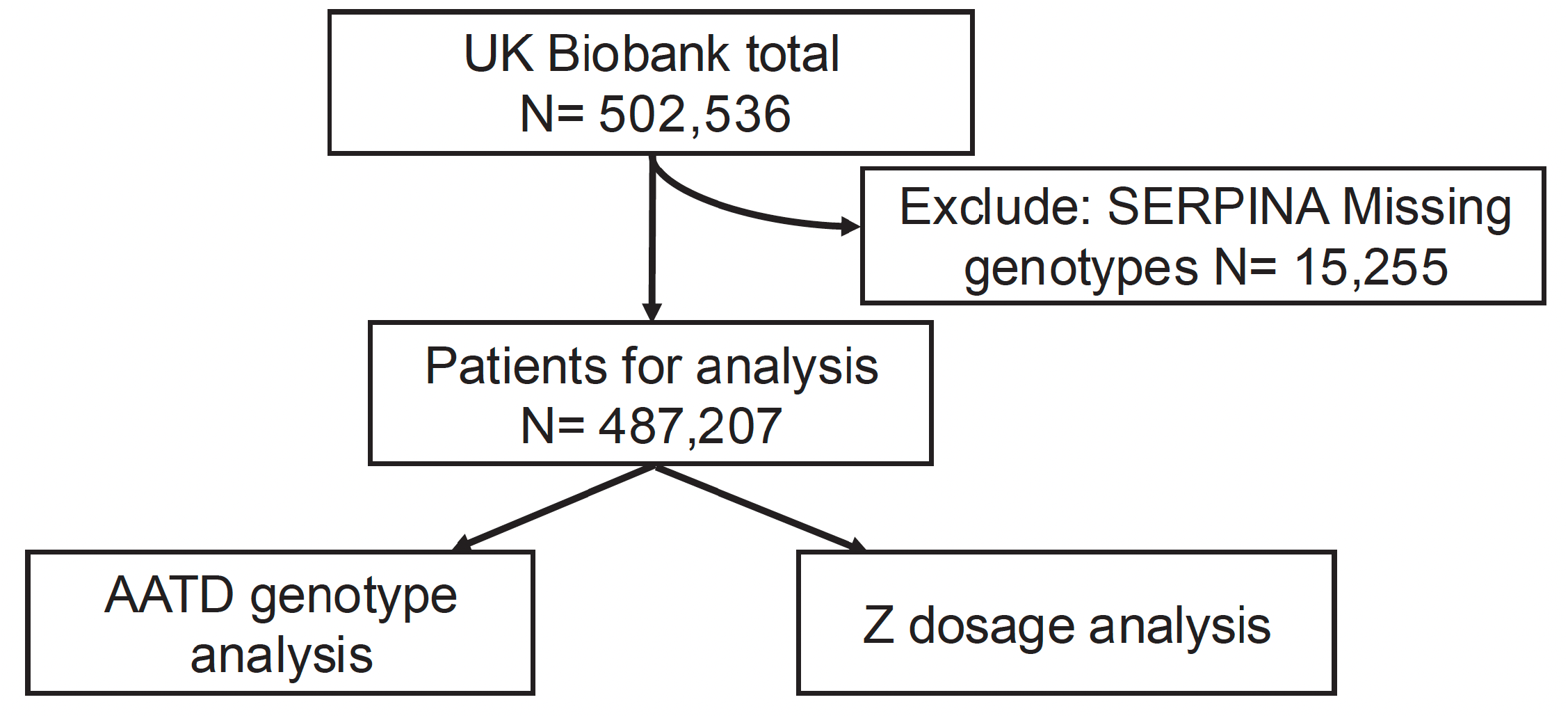
Flowchart of patient selection from the UK Biobank.

