## Supplementary Tables for "The Alpha-1 Antitrypsin Z-allele associates with sarcopenia in the UK Biobank"

**Supplementary Table 1. Full results of PheWAS analysis based on Z allele dosage**

| **Phenotype** | **n** | **Beta** | **Standard Error** | **P value** | **P value (bonferroni)** |
| --- | --- | --- | --- | --- | --- |
| Albumin | 425196 | 0.68106752 | 0.020957 | 2.15E-231 | 2.68E-227 |
| Globulin | 421090 | 5.13831157 | 0.222439 | 5.50E-118 | 6.86E-114 |
| Doctor diagnosed alpha 1 antitrypsin deficiency | 118597 | 4.22743602 | 0.233400 | 2.54E-73 | 3.17E-69 |
| Total protein | 424735 | 0.55641323 | 0.032497 | 1.08E-65 | 1.34E-61 |
| ALP | 464334 | 3.07580951 | 0.201738 | 1.79E-52 | 2.23E-48 |
| GGT | 425054 | 0.0114376 | 0.000752 | 3.18E-52 | 3.97E-48 |
| Standing height | 485769 | 1.00986464 | 0.068818 | 9.62E-49 | 1.20E-44 |
| ALT | 464137 | 1.12593813 | 0.108409 | 2.89E-25 | 3.61E-21 |
| Spirometry acceptability | 485338 | 0.37196718 | 0.035976 | 4.71E-25 | 5.88E-21 |
| Body fat percentage | 106046 | -0.2546454 | 0.024783 | 9.39E-25 | 1.17E-20 |
| Fat free mass | 106046 | -0.2657457 | 0.026119 | 2.65E-24 | 3.30E-20 |
| Whole body water mass | 106009 | -0.2597647 | 0.025933 | 1.32E-23 | 1.64E-19 |
| Basal metabolic rate | 106009 | -0.2458364 | 0.024654 | 2.08E-23 | 2.59E-19 |
| Body fat mass | 92681 | -0.2579879 | 0.027169 | 2.24E-21 | 2.79E-17 |
| Fat mass | 92681 | -0.2478172 | 0.026546 | 1.03E-20 | 1.28E-16 |
| AST | 462598 | 0.75501713 | 0.081661 | 2.35E-20 | 2.93E-16 |
| Lean mass | 91961 | -0.2456917 | 0.026922 | 7.25E-20 | 9.05E-16 |
| Skeletal muscle mass | 91961 | -0.2418519 | 0.027622 | 2.06E-18 | 2.57E-14 |
| Triglycerides | 420525 | 0.42263105 | 0.048856 | 5.14E-18 | 6.41E-14 |
| Total cholesterol | 464371 | -0.0006746 | 0.000078 | 5.15E-18 | 6.43E-14 |
| AST/ALT ratio | 464137 | -0.0283298 | 0.003310 | 1.14E-17 | 1.42E-13 |
| ALT/AST ratio | 462598 | 0.01822589 | 0.002192 | 9.22E-17 | 1.15E-12 |
| Platelet count | 419191 | -0.0512187 | 0.006850 | 7.59E-14 | 9.47E-10 |
| Urea | 464287 | 0.01004869 | 0.001345 | 8.07E-14 | 1.01E-09 |
| Ankle spacing width right | 159166 | 0.41049222 | 0.059862 | 7.04E-12 | 8.78E-08 |
| Ankle spacing width left | 159168 | 0.41495525 | 0.060573 | 7.39E-12 | 9.22E-08 |
| Heel bone mineral density | 274410 | 0.31025483 | 0.045326 | 7.66E-12 | 9.56E-08 |
| Arm lean mass DXA | 38598 | 20.4935153 | 3.019428 | 1.16E-11 | 1.45E-07 |
| Hemoglobin | 464372 | -0.0160795 | 0.002386 | 1.58E-11 | 1.97E-07 |
| Legs combined bone area | 39709 | 18.0540669 | 2.702420 | 2.41E-11 | 3.01E-07 |
| Legs bone area DXA | 39709 | 18.0540669 | 2.702420 | 2.41E-11 | 3.01E-07 |
| Leg lean mass | 38598 | 13.0045699 | 2.006707 | 9.25E-11 | 1.15E-06 |
| Impedance of arm right | 478404 | 2.56907625 | 0.415932 | 6.55E-10 | 8.18E-06 |
| Impedance of arm left | 478429 | 2.58200993 | 0.426820 | 1.45E-09 | 1.82E-05 |
| Arm lean mass | 38598 | 0.91467791 | 0.151187 | 1.46E-09 | 1.82E-05 |
| Impedance of whole body | 478414 | 3.98452216 | 0.667681 | 2.41E-09 | 3.00E-05 |
| WBC count | 461798 | -0.2589201 | 0.043583 | 2.84E-09 | 3.54E-05 |
| Reticulocyte percentage | 464371 | -0.0399011 | 0.006736 | 3.16E-09 | 3.94E-05 |
| Average weekly beer plus cider intake | 336890 | -0.2938808 | 0.049758 | 3.50E-09 | 4.37E-05 |
| Height | 47706 | 1.32694408 | 0.228273 | 6.18E-09 | 7.71E-05 |
| Trunk lean mass | 38591 | 0.39607715 | 0.068338 | 6.85E-09 | 8.55E-05 |
| Diastolic blood pressure | 449531 | -0.4602743 | 0.079608 | 7.40E-09 | 9.23E-05 |
| RBC count | 419191 | -0.0027275 | 0.000477 | 1.09E-08 | 0.000135452 |
| FVC best | 346230 | 0.04851629 | 0.008522 | 1.25E-08 | 0.000155598 |
| Reticulocyte count | 464371 | -0.0016726 | 0.000296 | 1.55E-08 | 0.000193426 |
| Creatinine (urine) | 464009 | 0.05921424 | 0.010681 | 2.96E-08 | 0.00036952 |
| Impedance of leg right | 478442 | 1.4970924 | 0.270095 | 2.98E-08 | 0.000371555 |
| Bone density | 274410 | -1.9220046 | 0.350754 | 4.27E-08 | 0.000532266 |
| FVC average | 443926 | 0.04334564 | 0.008189 | 1.20E-07 | 0.001499214 |
| Bilirubin | 394279 | 0.03692458 | 0.007009 | 1.38E-07 | 0.001722711 |
| Pelvis bone area | 39709 | 6.08292631 | 1.156819 | 1.46E-07 | 0.001823368 |
| Platelet count | 419191 | 0.4187483 | 0.079954 | 1.63E-07 | 0.002033728 |
| Systolic blood pressure | 449527 | -0.7562689 | 0.144430 | 1.64E-07 | 0.002045818 |
| Head bone area | 39709 | 2.57854252 | 0.502884 | 2.95E-07 | 0.003680787 |
| Trunk fat mass | 478149 | 0.19695426 | 0.038777 | 3.79E-07 | 0.004733187 |
| Impedance of leg left | 478436 | 1.35338091 | 0.267767 | 4.32E-07 | 0.005391488 |
| Doctor diagnosed emphysema | 118597 | 0.87426677 | 0.176660 | 7.46E-07 | 0.009314444 |
| Arms combined bone area | 39709 | 8.55333716 | 1.754865 | 1.10E-06 | 0.013693638 |
| Neutrophils | 485338 | 0.14887433 | 0.031138 | 1.74E-06 | 0.021751966 |
| C reactive protein | 463309 | -0.1582294 | 0.033291 | 2.01E-06 | 0.025024072 |
| Heel bone mineral density | 274273 | -0.0064244 | 0.001364 | 2.47E-06 | 0.030763806 |
| Physical activity | 442301 | 0.21235196 | 0.045224 | 2.66E-06 | 0.033193102 |
| Heel bone mineral density T score automated | 274410 | -0.0573233 | 0.012222 | 2.73E-06 | 0.034056919 |
| L1-L4 area BMD | 37804 | 1.06915786 | 0.228525 | 2.90E-06 | 0.036179551 |
| Leg bone area left | 14473 | 10.3670333 | 2.221329 | 3.08E-06 | 0.038470713 |
| Lean mass | 45316 | 21.035395 | 4.526106 | 3.37E-06 | 0.042027429 |
| Fat lean measure | 39463 | 0.01294366 | 0.002800 | 3.80E-06 | 0.047429293 |
| Leg bone area right | 14473 | 10.2452202 | 2.218258 | 3.90E-06 | 0.048625494 |
| Associations between Z allele dosage and all available phenotypes using linear regression for continuous outcomes and logistic regression for binary outcomes, adjusted for age, sex, smoking status, assessment center, and genetic principal components (PC1-PC5). Statistical significance was defined using a Bonferroni correction for multiple testing (p < 0.05 / N), where N is the number of phenotypes tested). | | | | | |

**Supplementary Table 2A – Measures of fat free mass index and appendicular skeletal muscle mass index by AAT genotype**

|  | **FFMI β coefficient (95% CI)** | **Adj FFMI β coefficient (95% CI)** | **Adj* FFMI β coefficient (95% CI)** | **ASMI β coefficient (95% CI)** | **Adj ASMI β coefficient (95% CI)** | **Adj* ASMI β coefficient (95% CI)** |
| --- | --- | --- | --- | --- | --- | --- |
| **MS** | −0.005 (95% CI −0.031, 0.021) | −0.007 (95% CI −0.025, 0.012) | −0.007 (95% CI −0.027 to 0.012) | −0.007 (95% CI −0.021 to 0.007) | −0.006 (95% CI −0.016 to 0.005) | −0.006 (95% CI −0.017 to 0.005) |
| **SS** | −0.040 (95% CI −0.200, 0.121) | −0.072 (95% CI −0.184, 0.040) | −0.051 (95% CI −0.171 to 0.070) | **−0.023 (95% CI −0.107 to 0.061),** | −0.039 (95% CI −0.099 to 0.022) | −0.028 (95% CI −0.093 to 0.037) |
| **MZ** | **−0.125 (95% CI −0.166, −0.085)** | **−0.107 (95% CI −0.135 to −0.079)** | **−0.115 (95% CI −0.145 to −0.085)** | **−0.066 (95% CI −0.087 to −0.045)** | **−0.054 (95% CI −0.069 to −0.038)** | **−0.058 (95% CI −0.074 to −0.041)** |
| **SZ** | **−0.201 (95% CI −0.377, −0.025)** | **−0.189 (95% CI −0.312 to −0.067)** | **−0.175 (95% CI −0.305 to -0.045)** | **−0.121 (95% CI −0.213 to −0.029)** | **−0.104 (95% CI −0.170 to −0.037)** | **-0.095 (95% CI −0.166 to -0.025)** |
| **ZZ** | −0.346 (95% CI −0.748, 0.056) | **−0.498 (95% CI −0.779 to −0.217)** | **−0.612 (95% CI −0.924 to −0.300)** | −0.111 (95% CI −0.321 to 0.100) | **−0.190 (95% CI −0.343 to −0.038)** | **−0.277 (95% CI −0.446 to -0.109)** |
| All models are versus MM and represent linear regression models.  Adjusted model represents a multivariate model adjusted for core covariates which include: age when attended assessment center, sex, smoking status (current / former / never), UK Biobank assessment center, and PCs 1-5. Ethnicity group removed because all ethnicity were White for certain AAT genotypes.  *Adjusted model includes core covariates, lung function (FEV1% predicted) and liver function (FIB4)  Bold = statistically significant | | | | | | |

**Supplementary Table 2B – Odds of FFMI and ASMI-defined sarcopenia by AAT genotype**

|  | **FFMI-defined sarcopenia OR (95% CI)** | **Adj FFMI-defined sarcopenia OR (95% CI)** | **Adj* FFMI-defined sarcopenia OR (95% CI)** | **ASMI-defined sarcopenia OR (95% CI)** | **Adj ASMI-defined sarcopenia OR (95% CI)** | **Adj* ASMI-defined sarcopenia OR (95% CI)** |
| --- | --- | --- | --- | --- | --- | --- |
| **MS** | **0.97** (95% CI 0.93–1.01) | 1.00 (95% CI 0.93 to 1.08) | 0.97 (95% CI: 0.93 to 1.02) | **1.01** (95% CI: 0.99 to 1.04) | 1.02 (95% CI: 0.99 to 1.05) | 1.01 (95% CI: 0.99 to 1.04) |
| **SS** | **0.98** (95% CI 0.77–1.24) | 1.21 (95% CI 0.82 to 1.79) | 0.96 (95% CI: 0.72 to 1.24) | **1.00** (95% CI: 0.86 to 1.16) | 1.00 (95% CI: 0.85 to 1.17) | 0.97 (95% CI: 0.81 to 1.16) |
| **MZ** | **1.19** **(95% CI 1.12–1.25)** | **1.11 (95% CI 1.00 to 1.23)** | **1.24** **(95% CI: 1.16 to 1.31)** | **1.13** **(95% CI: 1.09 to 1.17)** | **1.17 (95% CI: 1.12 to 1.22)** | **1.17 (95% CI: 1.13 to 1.23)** |
| **SZ** | **1.28** **(95% CI 1.00–1.61)** | 1.31 (95% CI 0.84 to 2.04) | **1.30** **(95% CI: 1.00 to 1.66)** | **1.02** (95% CI: 0.87 to 1.20) | 1.06 (95% CI: 0.89 to 1.26) | 1.01 (95% CI: 0.83 to 1.23) |
| **ZZ** | **2.24** **(95% CI 1.42–3.39)** | **3.04 (95% CI 1.37 to 6.76)** | **2.02** **(95% CI: 1.16 to 3.32)** | **1.52** **(95% CI: 1.08 to 2.11)** | **1.66 (95% CI: 1.14 to 2.41)** | **1.85 (95% CI: 1.20 to 2.80)** |
| All models are versus MM and represent logistic regression models.  Adjusted model represents a multivariate model adjusted for core covariates which include: age when attended assessment center, sex, smoking status (current / former / never), UK Biobank assessment center, and PCs 1-5. Ethnicity group removed because all ethnicity was White for certain AAT genotypes.  *Adjusted model includes core covariates, lung function (FEV1% predicted) and liver function (FIB4)  Bold = statistically significant | | | | | | |

**Supplementary Table 3A - Measures of fat free mass index and appendicular skeletal muscle mass index by Z allele dosage**

|  | **FFMI β coefficient (95% CI)** | **Adj FFMI β coefficient (95% CI)** | **Adj* FFMI β coefficient (95% CI)** | **ASMI β coefficient (95% CI)** | **Adj ASMI β coefficient (95% CI)** | **Adj* ASMI β coefficient (95% CI)** |
| --- | --- | --- | --- | --- | --- | --- |
| **Z dosage (1 allele)** | **-0.129 (-0.168 to -0.089)** | **-0.110 (-0.137 to -0.082)** | **-0.117 (-0.146 to**  **-0.088)** | **-0.068 (-0.089 to**  **-0.048)** | **-0.055 (-0.070 to**  **-0.041)** | **-0.059 (-0.075 to**  **-0.043)** |
| **Z dosage (2 alleles)** | -0.345 (-0.748 to 0.057) | **-0.497 (-0.778 to -0.216)** | **-0.610 (-0.922 to**  **-0.300)** | -0.110 (-0.321 to 0.101) | **-0.190 (-0.342 to**  **-0.037)** | **-0.277 (-0.445 to**  **-0.108)** |
| All models are versus MM and represent linear regression models.  Adjusted model represents a multivariate model adjusted for core covariates which include: age when attended assessment center, sex, smoking status (current / former / never), UK Biobank assessment center, and PCs 1-5. Ethnicity group removed because all ethnicity were White for certain AAT genotypes.  *Adjusted model includes core covariates, lung function (FEV1% predicted) and liver function (FIB4)  Bold = statistically significant | | | | | | |

**Supplementary Table 3B - Odds of FFMI and ASMI-defined sarcopenia by Z allele dosage**

|  | **FFMI-defined sarcopenia OR (95% CI)** | **Adj FFMI-defined sarcopenia OR (95% CI)** | **Adj* FFMI-defined sarcopenia OR (95% CI)** | **ASMI-defined sarcopenia OR (95% CI)** | **Adj ASMI-defined sarcopenia OR (95% CI)** | **Adj* ASMI-defined sarcopenia OR (95% CI)** |
| --- | --- | --- | --- | --- | --- | --- |
| **Z dosage (1 allele)** | **1.194 (1.131 to 1.260)** | **1.205 (1.141 to 1.273)** | **1.241 (1.169 to 1.316)** | **1.125 (1.086 to 1.165)** | **1.161 (1.117 to 1.206)** | **1.165 (1.117 to 1.215)** |
| **Z dosage (2 alleles)** | **2.246 (1.421 to 3.396)** | **2.532 (1.589 to 3.870)** | **2.028 (1.163 to 3.325)** | **1.517 (1.075 to 2.108)** | **1.661 (1.136 to 2.405)** | **1.846 (1.203 to 2.800)** |
| All models are versus MM and represent logistic regression models.  Adjusted model represents a multivariate model adjusted for core covariates which include: age when attended assessment center, sex, smoking status (current / former / never), UK Biobank assessment center, and PCs 1-5. Ethnicity group removed because all ethnicity was White for certain AAT genotypes.  *Adjusted model includes core covariates, lung function (FEV1% predicted) and liver function (FIB4)  Bold = statistically significant | | | | | | |

**Supplementary Table 4**

|  | **Combined FFMI and HGS -defined sarcopenia OR (95% CI)** | **Adj FFMI and HGS-defined sarcopenia OR (95% CI)** | **Adj* FFMI and HGS-defined sarcopenia OR (95% CI)** |
| --- | --- | --- | --- |
| **MS** | **0.881 (0.807 to 0.961)** | 0.930 (0.850 to 1.016) | 0.929 (0.780 to 1.099) |
| **SS** | 0.819 (0.448 to 1.355) | 0.885 (0.483 to 1.472) | 0.842 (0.258 to 2.003) |
| **MZ** | **1.150 (1.020 to 1.291)** | **1.218 (1.079 to 1.369)** | 1.072 (0.832 to 1.358) |
| **SZ** | **0**.777 (0.388 to 1.371) | 0.849 (0.422 to 1.505) | 1.693 (0.595 to 3.771) |
| **ZZ** | **3.463 (1.554 to 6.635)** | **4.108 (1.818 to 8.039)** | 3.722 (0.576 to 13.692) |
| All models are versus MM and represent logistic regression models.  Adjusted model represents a multivariate model adjusted for core covariates which include: age when attended assessment center, sex, smoking status (current / former / never), UK Biobank assessment center, and PCs 1-5. Ethnicity group removed because all ethnicity was White for certain AAT genotypes.  *Adjusted model includes core covariates, lung function (FEV1% predicted) and liver function (FIB4)  Bold = statistically significant | | | |

**Supplementary Table 5**

|  | **Combined FFMI and HGS -defined sarcopenia OR (95% CI)** | **Adj FFMI and HGS-defined sarcopenia OR (95% CI)** | **Adj* FFMI and HGS-defined sarcopenia OR (95% CI)** |
| --- | --- | --- | --- |
| **Z dosage (1 allele)** | **1.145 (1.018 to 1.282)** | **1.209 (1.073 to 1.356)** | 1.107 (0.867 to 1.391) |
| **Z dosage (2 alleles)** | **3.501 (1.571 to 6.709)** | **4.137 (1.830 to 8.094)** | 3.749 (0.580 to 13.789) |
| All models are versus MM and represent logistic regression models.  Adjusted model represents a multivariate model adjusted for core covariates which include: age when attended assessment center, sex, smoking status (current / former / never), UK Biobank assessment center, and PCs 1-5. Ethnicity group removed because all ethnicity was White for certain AAT genotypes.  *Adjusted model includes core covariates, lung function (FEV1% predicted) and liver function (FIB4)  Bold = statistically significant | | | |
