## Supplementary Methods for "The Alpha-1 Antitrypsin Z-allele associates with sarcopenia in the UK Biobank"

**Study Population and Data Source.** The UKBB is a large, population-based prospective cohort comprising data from 502,536 individuals aged 37–73 years who were recruited from across the United Kingdom and resided within 25 miles of 22 assessment centers located in England, Wales, and Scotland. At enrollment, participants completed health-related questionnaires via touchscreen interfaces or structured interviews, underwent standardized physical assessments, and contributed biological specimens. All participants provided written informed consent. Data access and analyses were performed following approval by the Institutional Review Board.

**Definitions for comorbidities**

When defining comorbidities, asthma was clinician-diagnosed or defined by having ICD9 codes 4930; 4931; 4939, or ICD10 codes J45; J46. COPD was defined as either low FEV1/FVC ratio <0.7, or a composite definition (FEV1/FVC ratio <0.70, a clinician diagnosis of emphysema, chronic bronchitis, or COPD, or ICD9 codes 4910; 4911; 4912; 4918; 4919; 4929; 496* or ICD10 codes J41* - J44*) (1). Bronchiectasis was either clinician-diagnosed or defined by ICD9 code 4949, or ICD10 code J47. Liver disease was defined by ICD9 codes 571.0–571.6, 070.2–070.6, 572.2/572.4/572.8, 571.8–571.9, 155.0, V42.7, or ICD10 codes K70, 72, 74, 76, B18, Z944, C22. AATD was defined by clinician diagnosis, ICD9 codes 27761, 27762 or ICD10 code E880 (which is a broader code for disorders of plasma-protein metabolism and may include plasminogen deficiency)(2). Liver cancer was defined by clinician diagnosis, ICD9 codes 1550-1552; 2308, or ICD10 codes C22*. Medications were self-reported (UKBB code 20003).

**Phenotypic data collection**

All phenotypic data were collected in accordance with standardized UKBB assessment protocols. Spirometric testing was performed with a Pneumotrac 6800 spirometer (Vitalograph Ltd., Buckingham, England) following standardized procedures. Percent predicted FEV1 was derived using Global Lung Initiative (GLI) reference equations(3). Respiratory symptoms were self-reported. Laboratory parameters included markers of systemic inflammation (CRP), liver function (AST, ALT), and lipid / lipoprotein metabolism (triglycerides). AST-to-Platelet Ratio Index (APRI) was computed as ("AST"/"ULN")/"platelet count"×100, using an upper limit of normal (ULN) for AST of 40 U/L. The AST/ALT ratio was calculated as AST divided by ALT. Fibrosis-4 index (FIB4) was calculated as ("age"x"AST")/("platelet count"x√("ALT" )), incorporating age (years), AST (U/L), ALT (U/L), and platelet count. High FIB4 was defined as ≥2.67(4), and high APRI was defined as ≥1.5(5).

Slow walking pace was determined by self-report. Body weight, fat mass, and fat-free mass were quantified using bioelectrical impedance analysis (BIA) with a Tanita BC-418 MA body composition analyzer (Tanita Europe). Participants removed shoes and heavy outer clothing prior to BIA assessment. Handgrip strength (HGS) was assessed using a Jamar J00105 hydraulic dynamometer. Measurements were obtained once from each hand following standardized instructions by trained staff, and the average of the dominant hand was used for handgrip strength in our analysis. For those who were ambidextrous, we took an average of the right- and left-hand values. Physical activity was evaluated by questionnaire items adapted from the validated short form of the International Physical Activity Questionnaire (IPAQ).
